# Acute care disruptions due to information technology failures in the Netherlands from 2000 to 2020

**DOI:** 10.1101/2023.02.13.23285875

**Authors:** Liselotte S. van Boven, Renske W.J. Kusters, Vincent W. Klokman, Christian Dameff, Dennis G. Barten

**Author notes:** **Corresponding author** D.G. Barten, Department of Emergency Medicine, VieCuri Medical Center, P.O. Box 1926, 5900 BX Venlo, The Netherlands. **Funding:** None. **Competing interests:** None. **Ethical approval** The institutional review board of VieCuri Medical Center approved the primary scoping review (#439).

## Abstract

**Background:** Healthcare organizations are becoming increasingly dependent on Information technology (IT) for the provision of routine services. IT failures within hospital systems can impact acute patient care, including emergency department (ED) closure and the cancellation of operations. This study aims to gain insight into the impact of hospital IT failures in the Netherlands on acute care delivery and to provide recommendations for future preparedness.

**Methods:** This is a systematic scoping review of major acute care disruptions in Dutch hospitals between 2000 and 2020. Incidence of etiology, duration, ED diversion, and closure of multiple locations was evaluated. IT failures were included when associated with the closure of hospital departments and/or evacuations.

**Results:** Thirty-nine IT failures resulting in acute care disruption were identified. The majority occurred between 2010 and 2020 (n=37, 95%). Of the 39 events, 33 (85%) were primary IT failures and were mainly caused by computer network and/or hospital software failure. Secondary events predominantly resulted from power failure. Most events (n= 36, 92%) were resolved within minutes to hours. All events were associated with an ED closure, 27 (69%) with an operating room (OR) stop and two (5%) with external hospital evacuation of one or more patients. Furthermore, 17 incidents (44%) involved multiple hospital locations, and seven (41%) of these involved closure of multiple locations with an ED.

**Conclusion:** The impact of IT failures on acute care disruptions in the Netherlands has considerably increased since 2010. This stresses the urge to improve IT security and business continuity in today’s hospitals.

**PUBLIC INTEREST SUMMARY:** As the healthcare industry is becoming increasingly digitalized, technological failure potentially has a major impact on hospitals and patient care continuity. The current study found that Information Technology (IT) failures in Dutch hospitals between 2000 and 2020 are increasingly associated with acute care disruptions. IT failures were mainly caused by software or hardware failures or by power outages. Preventive measures and emergency planning may lessen the impact and ensure improved business- and patient care continuity.

## INTRODUCTION

Healthcare organizations, hospitals in particular, are increasingly dependent on information technology (IT). IT-related functions involve hospital and pre-hospital emergency medical services (EMS) communication systems, radiology systems including imaging devices, electronic health records (EHR), and even robot-assisted surgery.(1)(2) The digital healthcare revolution has greatly improved efficiency and has helped to improve and optimize patient care and hospital staff productivity.(3) Conversely, failure or malfunction of IT systems has serious and widespread effects on hospital care continuity, including acute care service delivery.(4)(5) A recent analysis of internal hospital disasters and crises (IHCDs) in the Netherlands revealed an increasing downtime trend, likely attributed to IT failures.(4) Similarly, international downtime numbers are rising.(6)

Causes of IT disruptions are diverse and may include technical failures, power outages, and fires, as well as human-induced triggers such as planned downtime for system updates or cyberattacks.(1) A hardware malfunction in a Dutch academic hospital in 2018 that led to IT system failure with a resultant five-hour emergency department (ED) diversion demonstrates the potential and serious consequences of healthcare IT failure.(5) Downtime, whatever the cause, may compromise laboratory and radiology services, medication administration and reporting, and patient care handovers between clinical staff.(1) Consequently, inpatient stays may be prolonged and time-sensitive medical procedures may be delayed.(1)(7) The digitalization of the healthcare sector poses additional potential hazards. The more healthcare professionals rely on the availability of IT services, the more complicated it becomes to provide continuity of care when IT services are disrupted.(8) The ED is particularly vulnerable to this threat because of the high turnover of patients and the acuity of their complaints, combined with a high dependency on rapid test results and clinical data availability to ensure safe patient triage and treatment.(9) When multiple hospital locations are part of a larger healthcare organization, they may share the same IT systems, including a connected EHR. A failure of connected EHR systems poses the risk that acute care may be disrupted across multiple healthcare center locations encompassing a large geographical region. Such issues potentially endanger patient wellbeing as travel time to a fully operational hospital may be longer. Therefore, an adequate understanding of these incidents is essential and may lead to better preparedness and response in the future.

The limited medical literature on IT failures mainly focuses on downtime preparedness by hospital staff (10)(11) or hospital staff experiences regarding downtime.(9)(12)(13) However, little is known about the incidence of IT failures and their impact on the provision of acute care services. Therefore, the aim of this study was to identify and characterize major hospital IT failures in the Netherlands between 2000 and 2020, focusing on the impact of healthcare IT failure on continuity of acute care services.

## 2. METHODS

### 2.1 Study design

A systematic scoping review of acute care disruptions in the Netherlands between 2000 and 2020 was conducted, from which IT failures were extracted. The framework described by Arksey & O’Malley (14) was followed and the review was reported per PRISMA-ScR guidelines (15). The scoping review protocol was developed *a priori* to guarantee reproducibility and transparency. The scoping review methodology and its dataset are available upon request. The institutional review board of VieCuri Medical Center approved this research project (#439).

### 2.2 Search and article selection

The search terms “hospital,” “closed,” “ICU,” “ED,” “department,” and “failure” and their synonyms were combined using Boolean operators. The scoping review assessed news articles retrieved from the LexisNexis database, Google, Google News, PubMed, and EMBASE and included articles if one or more hospital departments or intervention units (such as operating rooms (ORs) or cardiac intervention units) of Dutch hospitals faced sudden and unexpected closure. Events in which inpatient or critical care departments were unaffected or if the organization was not a fully-fledged hospital (i.e., hospitals containing an ED and Intensive Care Unit (ICU)) were not included. Partial closures and partial evacuations were excluded.

### 2.3 Data extraction and analysis

Individual events regarding IT failures were extracted from the database of acute care disruptions. Extracted variables included: location, temporal aspects, failure types, casualties, departments involved, evacuation, patient presentation stops, injuries, damages, recovery method, primary or subsequent event, and internal disaster or combined internal and external disaster. The data were descriptively analyzed using mathematical summations based upon the data gathered in an Excel document (Microsoft ® Excel for Mac, version 16.62, 2021).

### 2.4 Definitions

For the scope of this study, an IT failure was defined as “any error in telephone, internet or computer-based systems”.(4) Computer-based failure was further categorized into software failure (i.e., computer programs run on hardware), hardware failure (i.e. physical components of a system, e.g., a processor or computer monitor), failure of the patient information system (i.e., functionalities of the EHR), and cyberattack (i.e., attack via cyberspace for the purpose of disrupting, disabling, destroying, or maliciously controlling a computing environment/infrastructure).(16)

## 3. RESULTS

Between 2000 and 2020, a total of 39 IT failures were associated with hospital department closure and/or the evacuation of patients. Two incidents of IT failure (5%) occurred between 2000 and 2010; the remaining 95% (37/39) occurred between 2010 and 2020 (Fig 1). Thirty-three incidents (85%) concerned a primary IT failure, whereas six IT failures (15%) resulted from a cascade of events (Fig 2). The primary IT failure events were caused by computer network and/or software failures (12/33, 36%), scheduled IT updates (4/33, 12%), hardware failures (3/33, 9%), and telephone system technical failure (3/33, 9%). The cause was unknown in 11 events (33%) (Fig 2). Three of the non-primary IT failures (50%) resulted from an external power failure; two (33%) resulted from technical failure followed by internal power problems and eventually IT failure; and one incident (17%) resulted from internal power failure, leading to IT failure (Fig 2). Failure of the IT back-up systems was reported in 9 incidents (23%).

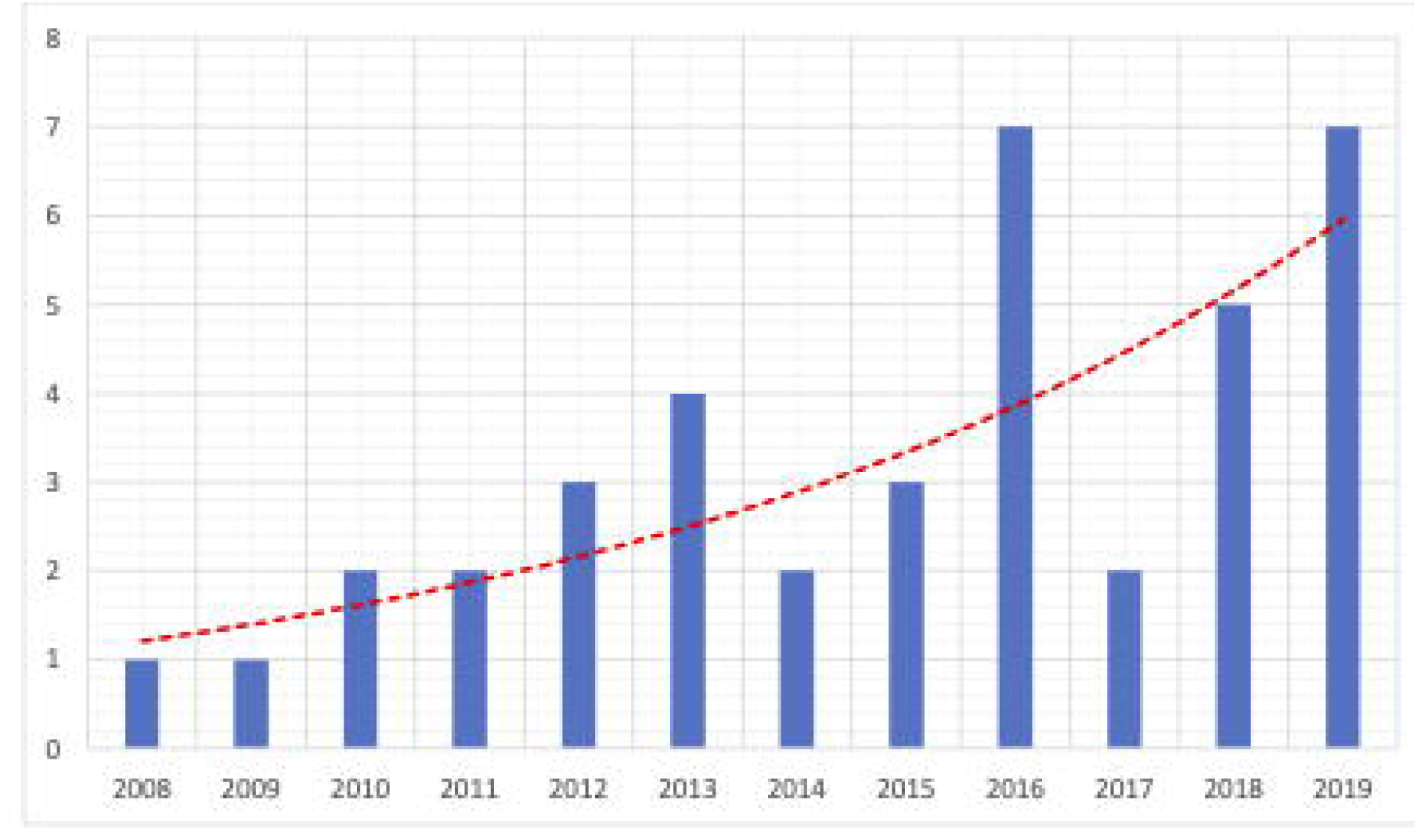

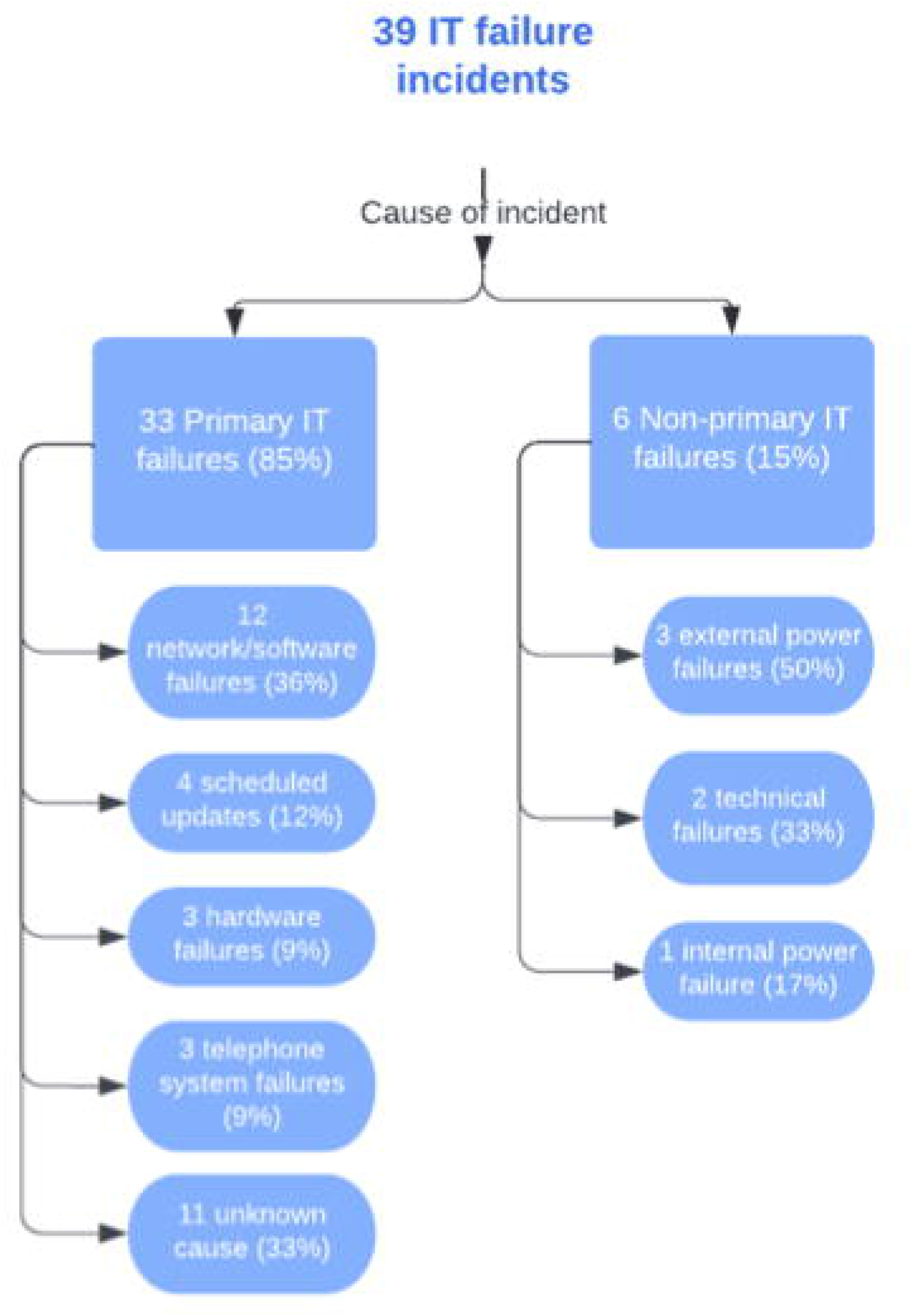

Most events involving IT failure lasted several minutes to hours and were resolved within one day (36/39, 92%). One event (3%) lasted for two days, one event lasted for four days, and one event lasted for five days before the system was functional again. Data regarding precise duration was not available for most events. In addition, 27 incidents (69%) took place during regular business hours, whereas the remaining 12 events (31%) took place in the evening or during the night. Out of all 39 incidents, four (10%) took place during the weekend (Saturday or Sunday).

In 43% of the incidents (17/39), multiple hospitals were affected at once, 49% (19/39) affected a single hospital and in 8% (3/39) the number of hospitals affected was unknown (Fig 3). In the cases where multiple healthcare centers were affected, many (7/17, 41%) involved the closure of multiple hospital locations with an ED (Fig 3).

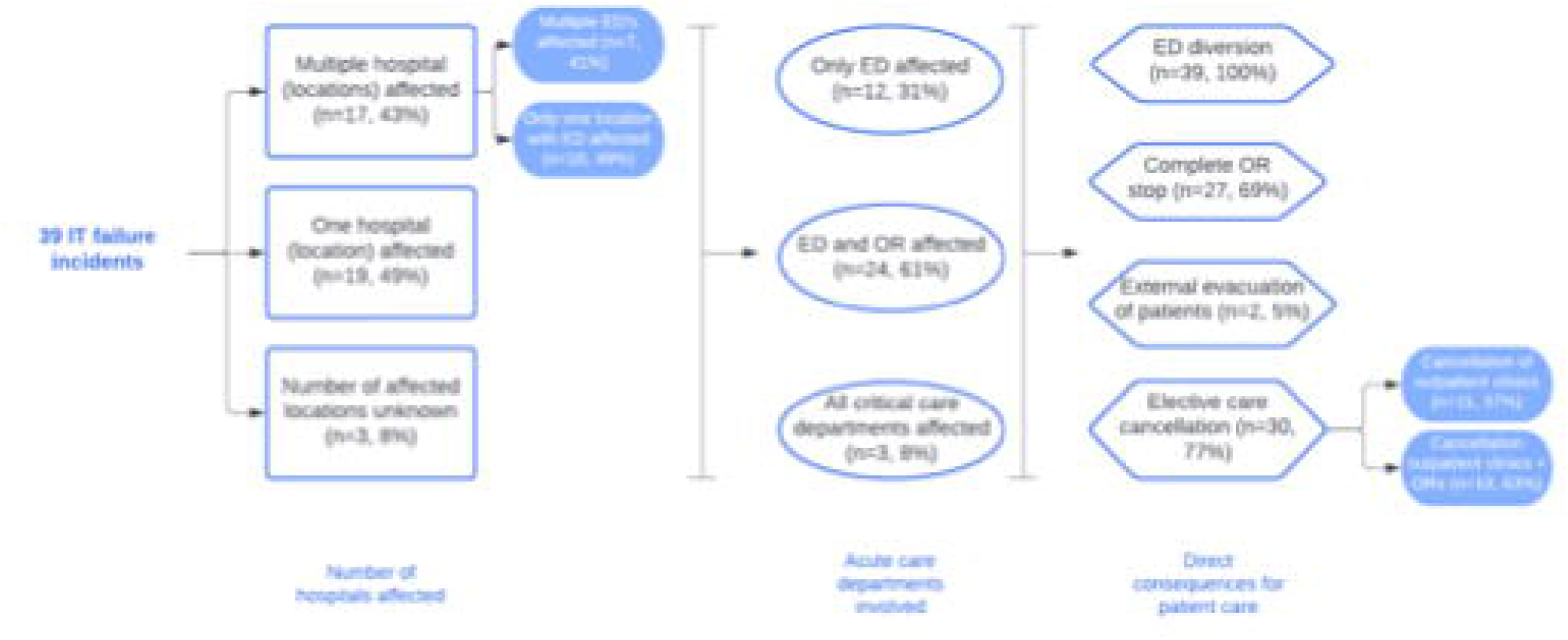

All IT failure incidents in this analysis resulted in an ED closure, 69% (27/39) included OR closures, and 5% (2/39) involved the external evacuation of patients (Fig 3). When focusing on acute care departments, 31% (12/39) only affected the ED, 61% (24/39) affected the ED and the OR, and 8% (3/39) affected all critical care departments (i.e., ED, OR, ICU, cardiac care unit) (Fig 3). Of all events, 77% (30/39) involved the cancellation of elective care; several involved the cancellation of outpatient clinics (11/30, 37%), and the remaining 63% (19/30) also involved cancellation of planned ORs (Fig 3). None of the news articles reported patient harm, and no events involved casualties.

## 4. DISCUSSION

Healthcare provision in hospitals is increasingly dependent on the proper functioning of hospital IT services. Therefore, IT failures can have a major impact on acute care delivery. In this study, IT failures in Dutch hospitals were increasingly associated with acute care disruptions since 2010. Furthermore, a substantial proportion of IT failures coincided with the closure of multiple hospital locations and/or EDs at once, potentially endangering the provision of acute care in a wider region. These findings underline the need for increased awareness and preparedness and the importance of securing IT continuity in today’s hospitals.

Although this topic receives scant attention in the medical literature, some aspects of IT failure and the impact on healthcare have already been assessed. Similar to our findings, a previous study investigating IT-related incidents in the United States (U.S.) has shown an increased incidence of IT failures in hospitals over time.(6) Twenty-one percent of incidents were caused by internal IT failure and problems with the EHR provider.(6) The majority of primary IT failures in our study had an internal cause (22/33, 67%) but no data were available regarding problems with the EHR provider. The same study by Larsen et al found that almost half of the analyzed incidents were due to cyberattacks, whereas cyberattacks were not among the causes for IT failure in our data set.(6) This inconsistency in findings may be explained by the fact that our study only included (major) IT failure incidents that were associated with the closure of one or more hospital departments or intervention units and/or evacuations. In the U.S. study, however, any incident regarding EHR downtime was included.(6) Like our study identified impacts of IT failures on multiple healthcare centers at once, Larsen et al stated that multiple hospitals were struck simultaneously, but no further details were provided. Consistent with our findings, a database study investigating patient safety reports of EHR downtime events in the U.S. found that IT downtime complicated the use of electronic patient files or temporarily rendered them unavailable.(1) The researchers also found that patient care was generally delayed and that viewing and uploading of radiology images was disrupted in the event of IT downtime.(1) This further emphasizes the potential for decreased patient wellbeing during downtime. Additionally, a hybrid study consisting of qualitative and quantitative analyses of downtime event records found a major impact of downtime on staff productivity, communication and perceived workload, and lab testing delays.(12)

The cause of primary IT failure incidents often involved internal causes; failures within the computer network and/or software as well as scheduled IT updates. This suggests that improvements from an IT department perspective may lead to a decreased incidence of IT failure events. Changes should include administrative attention, investment in cybersecurity, and optimal collaboration between IT and healthcare professionals.(2) Other key improvements include prioritization of processes and system applications, creation of an ‘IT emergency task force’, downtime planning for short- and long-term outages along with regular training scenarios.(2)(8) Of note, the cause of the IT failure remained unknown in a large number of incidents. This may be in part be due to the use of news and media outlets as data sources. Thorough root cause analysis of any event may improve preparedness for future events. The use of connected IT and/or EHR systems by multiple hospital (ED) locations may be another vulnerability as failure can lead to the closure of multiple hospital locations at once. Network segmentation, the division of a network into multiple smaller units, may reduce this risk.(17)

External causes of failure include external power failures. These may be avoided by the use of up-to-date and powerful back-up generators suitable for large facilities. (18) Another major external cause for healthcare IT failure concerns cyberattacks. Although cyberattacks were not among the causes of IT failures leading to acute care disruption in this cohort, there have been major incidents in other countries. These include the WannaCry ransomware attack on the National Health Service (NHS) in England in 2017 and the Conti attack on the Health Service Executive (HSE) in Ireland in 2021, in which hospitals reverted to a paper-based and manual system, outpatient appointments were cancelled, elective and day care procedures were postponed, and several EDs diverted ambulances to other care facilities.(19)(20) Prevention of cyberattacks by optimizing healthcare IT security is essential to avoid such significant impacts on patient care and wellbeing.(21) Such changes include early detection of suspicious digital activity,(22) segmentation of critical servers and applications (i.e., storing critical servers behind an additional firewall), and regular testing of the IT system.

Optimal hospital preparedness for events of IT failure is essential to ensure patient safety and continuity of care. Whereas most hospitals have an emergency plan in place in case of an IT-related incident, these plans should be thoroughly reviewed, updated, and routinely practiced.(8) Practicing downtime events and smoother operating transitions may lessen the need for ED diversion and thus enhance patient care continuity. Scenario-based training is not only useful for IT staff; healthcare providers would also benefit by gaining the necessary skills to ensure patient care continuity in the event of a hospital IT failure. The 2020 report by the Dutch Safety Council advised hospitals to regularly review the dependence of care on IT systems and to assess the possible patient safety risks associated with IT failure. In addition, they advise systematic analysis of any IT failure occurring at the facility, to share lessons learned, and to organize (tabletop) scenario trainings for multidisciplinary teams. Using contingency planning guides to determine hospital preparedness and identify possible weaknesses may prove useful in the future.(11) Awareness and preparedness on all fronts is needed to ensure optimal patient care during hospital IT downtime.

## 5. STRENGTHS AND LIMITATIONS

The main strength of this study is the inclusion of all IT failures in the Netherlands that involved a significant impact on acute care services as reported in news media between 2000 and 2020. The common finding of multiple hospital locations and/or ED closures simultaneously stressed the importance of securing IT continuity in modern healthcare, an aspect that has not been discussed in previous research.

The main limitation of the study is retrieval of data from online news articles. Hence, information of interest may be missing or may be misrepresented and bias toward sensational events may have occurred. In addition, IT failures without a disruption of acute care are not included in this study, leading to an underestimation of IT failure occurrence. However, larger incidents in which hospital functioning and patient care continuity were significantly impacted were deemed likely to be reported by the news, so the risk of missed major IT failures is considered low. Underreporting of events in the earlier years of the inclusion period may have occurred due to less availability of online news platforms. IT failures may not always lead to ED closure or ambulance diversion in other health systems, which should be considered when applying results to other countries. Lastly, only hospitals with acute care services were included in the study; other types of healthcare facilities were not evaluated. To give a detailed view of IT failure impacts on acute care, future research should include data regarding length of ED stay, missed or incorrect diagnoses, and number of postponed operations, among others.

## 6. CONCLUSION

The impact of IT failures on acute care disruptions in the Netherlands has considerably increased since 2010. Many IT failures were associated with the closure of multiple hospital locations and/or EDs at once, possibly endangering acute care in a wider region. These findings underline the importance of securing IT continuity in today’s hospitals. Recommendations include reviewing and updating of emergency plans, scenario-based training for IT downtime and raising staff awareness on this topic.

## Data Availability

All data produced in the present work are contained in the manuscript

## Abbreviations

IT: Information technology
ED: emergency department
EMS: emergency medical services
EHR: electronic health records
IHCDs: internal hospital disasters and crises
ICU: Intensive Care Unit

